# A Machine-Learning Approach for Predicting Depression Through Demographic and Socioeconomic Features

**DOI:** 10.1101/2022.11.13.22281677

**Authors:** Joseph Sun, Rory Liao, Mikhail Y. Shalaginov, Tingying Helen Zeng

**Affiliations:** Phillips Academy Andover, Andover, MA, 01810; Department of Computer Science and Engineering, Ohio State University, Columbus, OH 43210, USA; Department of Materials Science and Engineering, MIT, Cambridge, MA, USA; Divison of Career Education, Academy for Advanced Research, and Development (AARD), Cambridge, MA 02142, USA

**Keywords:** mental health, depression, machine learning, classification, decision tree, feature importance

## Abstract

According to the World Health Organization, over 300 million people worldwide are affected by major depressive disorder (MDD) [1]. Individuals battling this mental condition may exhibit symptoms including anxiety, fatigue, and self-harm, all of which severely affect well-being and quality of life. Current trends in social media and population behavior bring up an urgent need for health professionals to strengthen mental health resources, improve access and accurately diagnose depression [2]. To mitigate the disparate impact of depression on people of different social and racial groups, this study identifies factors that strongly correlate with the prevalence of depression in U.S. adults using health data from the 2019 pre-pandemic National Health Institute Survey (NHIS) [3]. In this study we trained a random forest model capable of performing a classification task on American-adults survey data with an accuracy of 98.7%. Our results conclude that age, education, income, and household demographics are the primary factors impacting mental health. Awareness of these mental health stressors may motivate medical professionals, institutions, and governments to identify more effectively the at-risk people and alleviate their potential suffering from MDD. By receiving adequate mental health services, Americans can improve their quality of life and form a more fulfilling society.

## I. Introduction

The most common mental disorder and the leading cause of over two-thirds of global suicides each year, major depressive disorder (MDD) often goes undiagnosed and untreated [1]. Nearly 20% of Americans will experience a major depressive episode in their lifetime, an occurrence that lasts on average 27 weeks and has intensified during the COVID-19 pandemic [4, 5, 6]. The National Institute of Mental Health (NIMH) defines depression as the daily presence of the symptoms including sadness, anxiety, irritability, feelings of hopelessness and guilt, decreased appetite and fatigue, and suicidal thoughts, for at least two weeks [2]. Though all of these may be signs of depression, not all are required for a diagnosis.

Currently, most medical diagnoses of depression rely on patients reaching out to their health care providers. Depression is diagnosed qualitatively, and patients are then referred to mental health professionals like psychologists and psychiatrists. Due to the stigma associated with depression and the wide breadth of symptoms, many depressive symptoms go undiagnosed and untreated [2, 4]. Therefore, it is important to identify mental health stressors and populations most at-risk for depression, to improve diagnostic strategies.

Studies have shown that individuals with fewer financial assets and greater stress demonstrate higher risk for depression [7]. Similarly, education level, age, and marital status have been identified as important features in mental health [8]. Furthermore, the emerging field of neurourbanism focuses on the impact of urban environments on happiness and the risk of stress and depression to individuals [9, 10]. Machine learning has recently become an effective tool for studying the impact of these factors on mental health.

Previous studies have utilized ML algorithms such as Catboost, Logistic Regression, Naïve Bayes, Random Forest (RF), and Support Vector Machine (SVM) to predict depression in individuals [8, 11, 12]. Priya et al. found the highest accuracy with Catboost (82%) on 470 participants, while Haque et al. found the highest accuracy with a Random Forest (95%) model tested on 6000 participants in a national Australian health survey [11, 12]. Though there have been studies conducted on the relationship between specific health attributes and depression, there has not been a comprehensive, national U.S. study on depression to date. For this study, we chose a pre-pandemic health survey to avoid possible influence from the COVID-19 pandemic. Using the 2019 National Health Institute Survey (NHIS), we applied several machine learning models to classify depression.

## II. Methods and Materials

### A. Data preprocessing and partitioning

Raw data from the 2019 NHIS, provided by the Centers for Disease Control and Prevention (CDC) and available to the public, was imported into Python. This survey contains responses from 31,997 U.S. adults interviewed on 534 questions, which is the most comprehensive pre-pandemic national health survey. American adults living in the 50 states and the District of Columbia were cluster-sampled by geographic location, those with no fixed home address, active-duty personnel living abroad, and those living in long-term care institutions are not surveyed. Conducted in a face-to-face interview format, the survey includes many questions ranging from demographic info to chronic health conditions.

All data contain quantitative answers, with binary answers for Yes/No questions. The Pandas and NumPy Python packages were used to preprocess the data. First, features that contained any missing values were removed. Those features only belonged to a small number of sampled adults and it would have been difficult to replace or input them into the algorithm. Since many of these asked very specific questions about chronic illnesses, which was not the focus of this research, they were also removed without a serious impact to our demographic analysis. Ultimately, the processed dataset has been cleaned out and contains 163 features.

One feature in the dataset, titled “DEPEV_A: “Have you ever had depression?” was selected as the label for supervised machine learning classification. To this question there were given four different answers: “Yes,” “No,” “Refused,” and “Don’t Know.” 25 instances containing “Refused” and 38 instances containing “Don’t Know” were removed from the entire dataset, converging to a survey matrix of 31,934 (instances/people) × 163 (features/questions). Of all the respondents, 17% of adults reported that they had felt depressed sometime in their lifetime; 83% reported they did not feel depressed.

After browsing classification model leaderboards on the ‘Papers With Code’ database, four different models were chosen for training: 1) Catboost [13]; 2) K-Nearest Neighbors (KNN) [14]; 3) Random Forest (RF) [15]; and 4) XGBoost [16]. For each model, the data was randomly split into an 80:20 ratio for training and testing purposes. The Scikit-Learn module was used for partitioning, model training, and measuring performance metrics.

### B. Performance metrics

Three performance metrics were calculated for each classifier: (1) accuracy score, (2) F1 Statistic, and (3) Cohen’s Kappa. The accuracy was measured as the normalized number of truly predicted values, using the following abbreviations: TP = true positive, FP = false positive, TN = true negative, and FN = false negative [16].

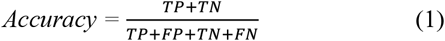

The F1 score is defined as the harmonic mean of two values, precision and recall [11]:

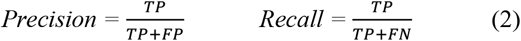

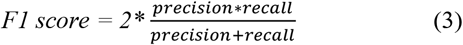

The third and final score used in this study is Cohen’s Kappa, which is expressed through the formula:

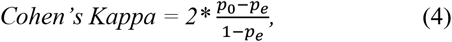

where p_0_ is the observed probability and p_e_ is the expected probability of the model prediction. A Cohen’s Kappa score of 1.0 is perfect agreement, while a score of 0.0 is considered no agreement [17].

### C. Preprocessing for feature importance

To evaluate feature importance, the features were combined into 22 feature categories and 1 as a label feature, to eliminate confounding variables (e.g., “Have you ever had diabetes?” and “Have you ever had prediabetes?”) that would decrease model accuracy. Highly correlated features, such as income-based questions, were filtered out as well. The heatmap in Fig. 1 shows the Spearman correlation coefficients between the remaining features. Due to the nature of this dataset, Spearman correlation was selected over Pearson correlation since it measures the strength and direction of monotonic relationships of ranked variables rather than linear association.

**Fig. 1.**
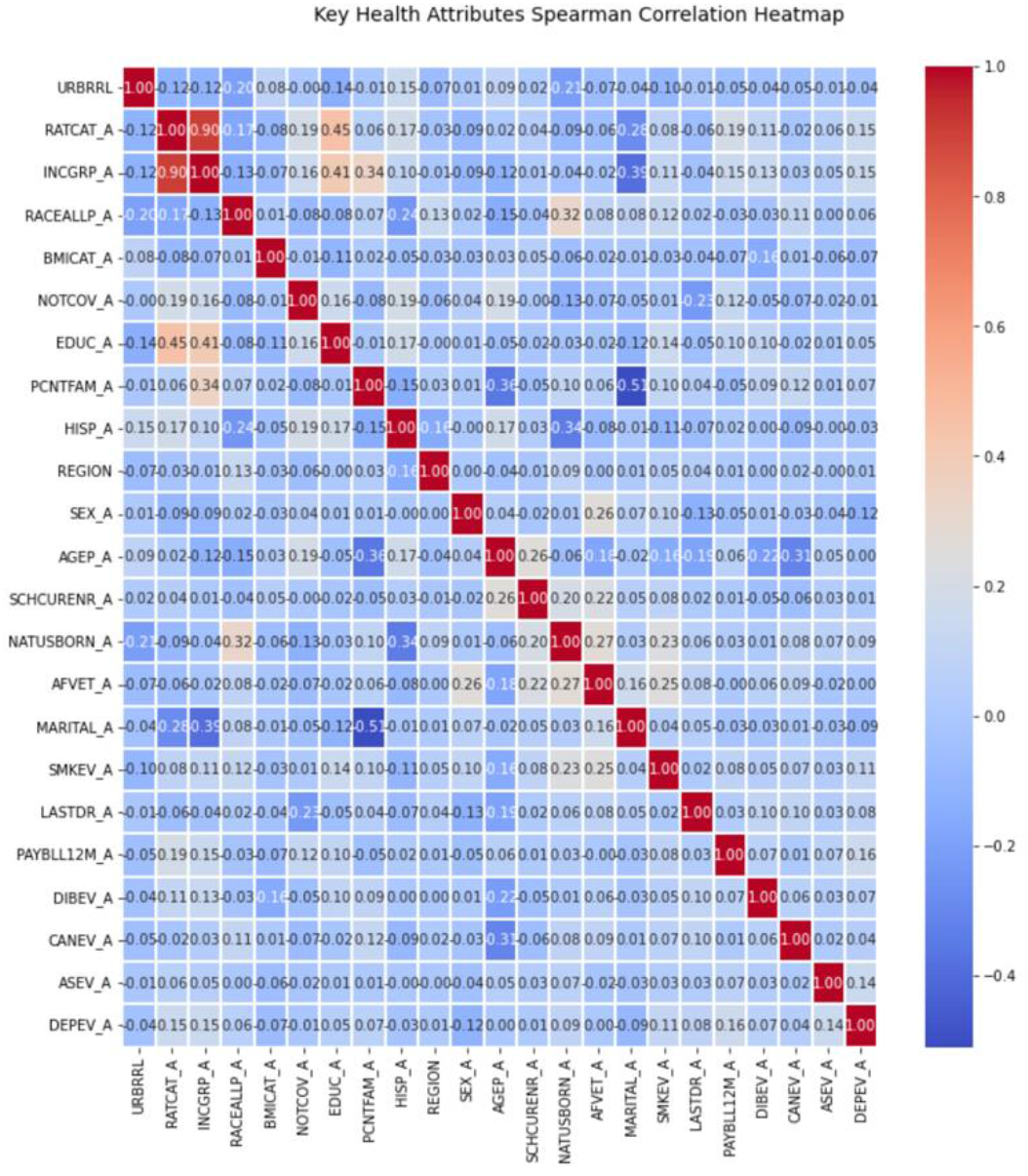
Spearman cross correlations heatmap of 23 selected demographic variables and health conditions including the label column “DEPEV: Have you ever had depression?” See [3] for detailed feature label information.

## III. Results and Discussion

### A. Comparison of models

Initially, the classification task was performed on the dataset of 162 features. Random Forest yielded the best results in all three performance categories, as shown in Table 1 below.

**TABLE I.** Evaluation of 4 Classification Models

| Model | Key metrics on 162 features for depression |  |  |
| --- | --- | --- | --- |
|  | Accuracy (%) | Weighted F1-Score | Cohen's Kappa |
| Catboost | 93.3 | 0.931 | 0.748 |
| KNN | 87.6 | 0.853 | 0.404 |
| Random Forest | **98.7** | **0.987** | **0.953** |
| XGBoost | 92.9 | 0.927 | 0.740 |

**TABLE II.** Random Forest Features Importance

| Variable Name | Top 5 Features |  |  |
| --- | --- | --- | --- |
|  | Description | Feature Score | Spearman's Correlation |
| AGEP_A | Age of sample adult | 0.190 | 0.002 |
| EDUC_A | Educational level of sample adult | 0.107 | 0.047 |
| RATCAT_A | Ratio of family income to poverty threshold | 0.010 | 0.145 |
| REGION | Household region within the U.S. | 0.073 | 0.006 |
| URBRRL | 2013 urban-rural classification scheme for counties | 0.070 | -0.044 |

Our Random Forest Classifier with repeated K-fold cross-validation performed the best with 100 trees, 10 splits, and 3 repeats. In cross validation sampling, RF was composed of an ensemble of decision trees, each sampling a portion of data without replacement. In a classification algorithm, each tree makes a prediction on the data and the classification takes the highest predicted class label across all trees. Since each decision tree only works with a subset of the total features (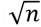 where n is the total number of features), the result from each tree varies and is not biased by influential features. Several repeats were used to reduce noise and variability from the performance. Thus, this model performs the best due to its efficiency and accuracy.

A final accuracy score of 98.7% is excellent, proving that health features of patients can provide very accurate predictions of depression through ML models. The confusion matrix in Fig. 2 shows the predictions.

**Fig. 2.**
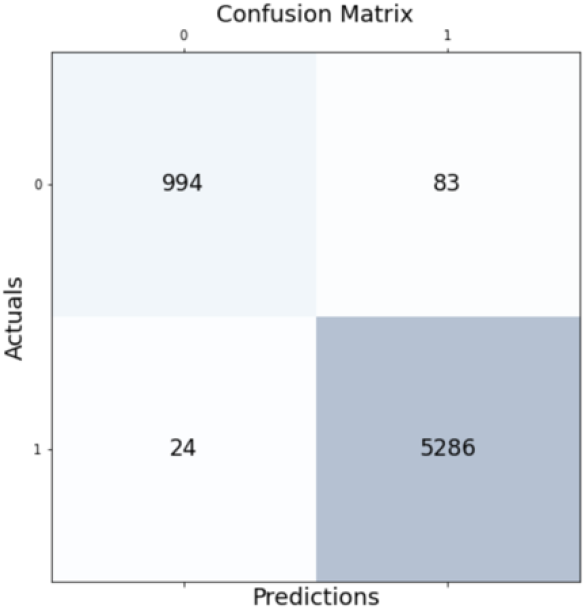
Confusion matrix of Random Forest classifier predictions versus actual values for depression in testing data. (0, 0) – true negative, (1, 1) – true positive, (0, 1) – false negative, and (1, 0) – false positive.

The two booster models, Catboost and XGBoost, also performed comparably well, with accuracies of 93.3% and 92.6%, respectively. A gradient boosting algorithm is particularly effective at improving accuracy by sequentially training decision trees to reduce the algorithm’s loss function. The Catboost model was trained with 10 iterations and a learning rate of 0.1. The XGBoost model was trained with the same learning rate and a maximum tree depth of 3.

Next, before feature importance analysis, the Random Forest model was retrained with only 22 key characteristics. The model performed well with an accuracy of 96.7%, an F1 score of 0.966, and a Cohen’s Kappa score of 0.876. The model performed slightly worse on the isolated characteristics because confounding variables that had high correlations with depression (e.g., “ANXEV – ever had depression”) were removed. With a smaller number of variables yet still high accuracy, however, this modeling can potentially be used to predict depression through health screenings. Though health practitioners and therapists should consult with their patients, this model may be useful for quantitatively confirming diagnoses of MDD.

### B. Feature importance

We hypothesized that certain demographic and socioeconomic features in different survey respondents would have pronounced effects on their likelihood of becoming depressed. Features importance were thus evaluated using all four models, but the RF algorithm was chosen as the most accurate predictor. In Fig. 3, the 22 features are ranked by importance, from most important to least.

**Fig. 3.**
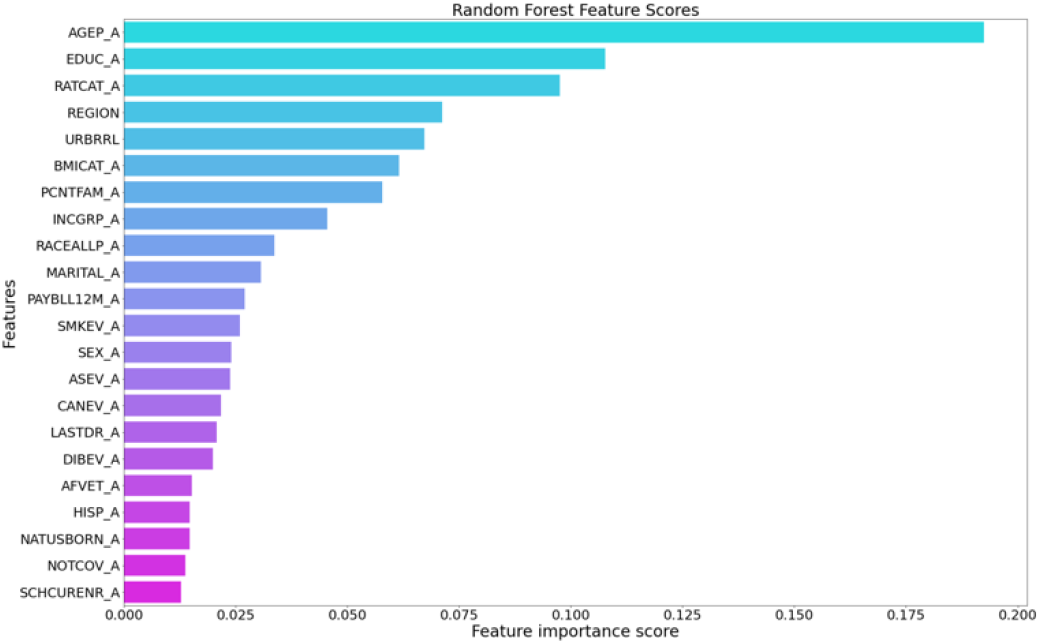
Graph of 22 features importance extracted from Random Forest classification model. The top five features include age, education, income, and geographic demographics. See [3] for detailed feature label information.

These features were then compared with the Spearman correlation coefficients to determine the direction of correlation.

In this survey, the answer choice of “Yes” was labeled “1” and the answer choice “No” was labeled “2.” Thus, a positive correlation coefficient corresponds with a lower prevalence of depression as the feature value increases. Note that Spearman correlation performs best with monotonic relationships, so it may be less accurate for features that vary. Nevertheless, it identifies the direction of correlation for our purposes.

The impact of these features on mental health is supported by previous literature. The feature of the highest importance, age, has consistently been noted in mental health research. Younger adults and adolescents have been identified as more at-risk for depression, due to a variety of reasons including puberty, school, and stress [18]. This is supported by the positive correlation—as age increases, depression prevalence decreases. Interestingly, however, it has the smallest correlation coefficient magnitude of all the variables above, at 0.003.

Not surprisingly, education level shows the second highest level of importance. The responses range from 00 (never attended kindergarten) to 11 (doctoral degree). It is reasonable that higher education can improve mental health, as indicated by the positive Spearman correlation, as it improves cognitive abilities, leading to more informed decisions, habits, and resources. Thus, when evaluating patients for depression, physicians should be aware that younger age and lower education level may be greater risk for depression. Furthermore, young adults studying in college and graduate schools have an even greater risk for stress and anxiety, putting them further at risk for depression.

“RATCAT_A” is an indicator for wealth and financial stability. A positive Pearson’s correlation coefficient of 0.149 indicates that a greater ratio of family income to poverty level— or greater wealth—is correlated with a smaller prevalence of depression. This supports recent findings in previous literature [8]. Greater wealth decreases stress from employment, paying bills, and can increase time for recreational activity and spending. This suggests the need for stronger mental health resources in lower-income communities to mitigate the effects of financial, employment, and familial stress.

Finally, both “REGION” and “URBRRL” denote household demographics. The regions are divided into four: Northeast, Midwest, South, and West. Though these values are not ranked in any particular order, the Spearman coefficient indicates that certain regions may show broader trends of mental health conditions across the U.S., which should be investigated further. “URBRRL” is ranked by four categories 1-4 where 1 denotes “large central metro” and 4 denotes “nonmetropolitan.” Thus, a negative correlation shows that depression is associated with more rural environments, while urban environments have a smaller prevalence of depression. This contrasts with previous studies on urbanism and depression, which conclude that depression is more prevalent in urban environments [9]. Due to the density of businesses and stimulating activity in the city, inhabitants are often more vulnerable to anxiety and stress; however, there are more available healthcare resources as well. These results indicate that there may be a lack of mental health counseling and resources in areas that are more rural.

### C. Patient Health Questionnaire (PHQ-8)

The Patient Health Questionnaire is the most widespread and reliable tool for diagnostic screening of MDD [4]. It consists of 8 questions concerning depressive symptoms, and assigns a value (0, 1, 2, 3) based upon the frequency of symptoms experienced.

Answers to these questions were included in the original dataset. Thus, these eight questions were isolated, and the Random Forest model once again provided the highest accuracy of 89.0%. More ML algorithms should be tested on PHQ data to improve accuracy, as it is a key tool to depression diagnosis.

## IV. Conclusions

In this study, we conclude that the Random Forest model as well as other decision tree and booster models are effective for predicting depression through health and demographic data. The attributes that most affect mental health are age, education, income, and geographical location. More in-depth neural networks may be applied to identify not only a binary value of depressed or nondepressed, but also the severity level of MDD in patients.

We propose that Twitter, Reddit, and social media should be used in future research into mental health, as it has a broad reach with people of all demographics, socioeconomic statuses, and health conditions. Feature analyses and sentiment analyses through Natural Language Processing, for example, may be conducted to predict severity of depression in online users. In addition, it would be important to analyze the pandemic’s effect on mental health stressors for the overall population in the past two years.

By identifying the key characteristics that affect populations at risk for depression, health professionals will have another diagnostic tool to aid them in making informed decisions regarding individual mental health treatments.

## Data Availability

All data produced in the present study are available upon reasonable request to the authors

## Acknowledgment

J.S. is grateful for the intern research opportunity offered by the Academy for Advanced Research and Development (AARD). This project is partially supported by the Scholarship of AARD for Future Scholars (http://www.ardacademy.org).

## Notes

### Competing Interest Statement

The authors have declared no competing interest.

### Funding Statement

This study was funded by the Academy for Advanced Research and Development (AARD).

## References

[1] Chiong, R., Budhi, G. S., Dhakal, S., & Chiong, F. (2021). A textual-based featuring approach for depression detection using machine learning classifiers and social media texts. Computers in Biology and Medicine, 135, 104499. https://doi.org/10.1016/j.compbiomed.2021.104499

[2] Depression. (n.d.). National Institute of Mental Health (NIMH). Retrieved August 13, 2022, from https://www.nimh.nih.gov/health/topics/depression

[3] NHIS – 2019 NHIS. (2021, April 5). https://www.cdc.gov/nchs/nhis/2019nhis.htm

[4] Costantini, L., Pasquarella, C., Odone, A., Colucci, M. E., Costanza, A., Serafini, G., Aguglia, A., Belvederi Murri, M., Brakoulias, V., Amore, M., Ghaemi, S. N., & Amerio, A. (2021). Screening for depression in primary care with Patient Health Questionnaire-9 (PHQ-9): A systematic review. Journal of Affective Disorders, 279, 473–483. https://doi.org/10.1016/j.jad.2020.09.131

[5] Miller, L., & Campo, J. V. (2021). Depression in adolescents. New England Journal of Medicine, 385(5), 445–449. https://doi.org/10.1056/NEJMra2033475

[6] Khubchandani, J., Sharma, S., Webb, F. J., Wiblishauser, M. J., & Bowman, S. L. (2021). Post-lockdown depression and anxiety in the USA during the COVID-19 pandemic. Journal of Public Health, 43(2), 246–253. https://doi.org/10.1093/pubmed/fdaa250

[7] Ettman, C. K., Abdalla, S. M., Cohen, G. H., Sampson, L., Vivier, P. M., & Galea, S. (2021). Low assets and financial stressors associated with higher depression during COVID-19 in a nationally representative sample of US adults. J Epidemiol Community Health, 75(6), 501–508. https://doi.org/10.1136/jech-2020-215213

[8] Lee, C., & Kim, H. (2022). Machine learning-based predictive modeling of depression in hypertensive populations. PLOS ONE, 17(7), e0272330. https://doi.org/10.1371/journal.pone.0272330

[9] Pykett, J. (2022). Why is emotional data failing to produce more humane cities? Urban governance and the (Interdisciplinary) problem of wellbeing. Urban Geography, 0(0), 1–19. https://doi.org/10.1080/02723638.2021.2003589

[10] Su, L., Zhou, S., Kwan, M.-P., Chai, Y., & Zhang, X. (2022). The impact of immediate urban environments on people’s momentary happiness. Urban Studies, 59(1), 140–160. https://doi.org/10.1177/0042098020986499

[11] Priya, A., Garg, S., & Tigga, N. P. (2020). Predicting anxiety, depression and stress in modern life using machine learning algorithms. Procedia Computer Science, 167, 1258–1267. https://doi.org/10.1016/j.procs.2020.03.442

[12] Haque, U. M., Kabir, E., & Khanam, R. (2021). Detection of child depression using machine learning methods. PLOS ONE, 16(12), e0261131. https://doi.org/10.1371/journal.pone.0261131

[13] Catboost classifier in python. (n.d.). Retrieved August 16, 2022, from https://kaggle.com/code/prashant111/catboost-classifier-in-python

[14] K nearest neighbor classification algorithm | knn in python. (2021, January 20). Analytics Vidhya. https://www.analyticsvidhya.com/blog/2021/01/a-quick-introduction-to-k-nearest-neighbor-knn-classification-using-python/

[15] Brownlee, J. (2020, April 19). How to develop a random forest ensemble in python. Machine Learning Mastery. https://machinelearningmastery.com/random-forest-ensemble-in-python/

[16] Brownlee, J. (2016, August 18). How to develop your first xgboost model in python. Machine Learning Mastery. https://machinelearningmastery.com/develop-first-xgboost-model-python-scikit-learn/

[17] Viera, A. J., & Garrett, J. M. (2005). Understanding interobserver agreement: The kappa statistic. Family Medicine, 37(5), 360–363.

[18] Kim, K. M. (2021). What makes adolescents psychologically distressed? Life events as risk factors for depression and suicide. European Child & Adolescent Psychiatry, 30(3), 359–367. https://doi.org/10.1007/s00787-020-01520-9

